# EXAMINING THE ROLE OF NON-SURGICAL THERAPY IN THE TREATMENT OF GERIATRIC INCONTINENCE

**DOI:** 10.1101/2021.10.21.21265353

**Authors:** Candace Parker-Autry, Rebecca Neiberg, X. Iris Leng, Catherine A. Matthews, Chantale Dumoulin, George Kuchel, Stephen B. Kritchevsky

**Affiliations:** Department of Urology, Obstetrics and Gynecology, Atrium Health Wake Forest Baptist, Winston-Salem, North Carolina, UNITED STATES; Biostatistics and Data Science, Wake Forest School of Medicine, Winston-Salem, North Carolina, UNITED STATES; School of Rehabilitation, University of Montreal, Montreal CANADA; UConn Center on Aging, University of Connecticut School of Medicine, Farmington, Connecticut, UNITED STATES; Gerontology and Geriatric Medicine, Wake Forest School of Medicine, Winston-Salem, North Carolina, UNITED STATES

## Abstract

**Objective:** To examine the role of physical function impairments on the change in urinary incontinence symptoms after pelvic floor muscle training in older women.

**Methods:** This is a prospective cohort study of 70 community-dwelling women, older than 70 years, with at least moderate incontinence symptoms. A comprehensive assessment of pelvic floor and physical function was done at baseline. Individualized PFM training prescriptions with behavioral management strategies to reduce incontinence episodes were provided for 12 weeks. Baseline physical function was determined using the Short Physical Performance Battery (total score of ≤9/12 defined impairment). A 3-day bladder diary established daily incontinence episodes. The change in urinary incontinence episodes based on presence of physical function impairment was our primary outcome. Descriptive analyses compared important demographic and clinical characteristics. Longitudinal mixed model linear regression analyses were used to assess for change in incontinence and estimate of improvement based on the presence of physical function impairment adjusted for age, race, and BMI.

**Results:** Participants’ mean±SD age was 76.9±5.4 years and 15.7% were African American with no significant differences in age or race between groups. Women with physical function impairment had higher mean ± SD BMI (33.6±14.5 vs 27.4±5.8 kg/m^2^; p=0.03) and significantly greater baseline incontinence episodes (4.5±2.9 vs. 2.7±2.1 episodes/day; p=0.005) than in women without functional impairment, respectively. After 6 weeks of pelvic floor exercises, women with physical functional impairment had a non-significant decrease in incontinence episodes/day compared to women with normal physical function (mean [95%CI], −1.2 [−2.0,−0.5] vs −0.4 [−1.1, 0.3], p=0.31). Women with physical function impairments also reported lower improvement in their incontinence symptoms compared to women without functional impairment (50.7±5.9% vs 64.2±5.9%, p=0.08).

**Conclusions:** Non-surgical therapy to include pelvic floor muscle training may not significantly decrease urinary incontinence symptoms to a degree that is satisfactory in women older than 70 years seeking treatment for urinary incontinence.

## INTRODUCTION

Urinary incontinence is a prevalent pelvic floor condition presenting in up to 50% of adult women. However, with aging beyond 70 years, urinary incontinence becomes heterogeneous, often evolving into a multi-factorial geriatric syndrome. Geriatric syndromes are defined as prevalent conditions present in older adults with shared risk factors such as physical function impairments, mobility disability, and cognitive decline.[1] Urinary incontinence and impaired physical function are inter-related geriatric conditions that result, in part, from skeletal muscle dysfunction with aging. Growing evidence supports that physical function impairment is a cause and consequence of urinary incontinence in aging adults.[2, 3]

In the 1940s, pelvic floor muscle training was developed to treat incontinence in young post-partum women under the premise that improved skeletal muscle bulk and functional efficiency of the pelvic floor and urethra sphincter would improve urethral closure and bladder support. Currently, pelvic floor muscle training is the crux of first-line therapy for treatment of urinary incontinence in women, regardless of age, based on data suggesting significant reductions in daily incontinence episodes after 6 or 12 weeks of training. [4] However, as women age beyond 70 years and develop UI symptoms, there is higher risk of concomitant development of skeletal muscle weakness and subsequent impairments in physical performance to include mobility disability and falls.[1] Impairments in physical function may decrease enrollment into clinical trials as there is increased concern for adverse events.[5] Further, women older than 70 years often have more severe and refractory urinary incontinence symptoms. [6] To date, providers treating geriatric incontinence have not considered the potential broader impact of aging-related changes in skeletal muscle health on urinary incontinence symptoms.

It is logical that healthy skeletal muscle structure and function are imperative to success of pelvic floor muscle training. With aging beyond 70 years, up to 60% of older women will live with physical function impairments and mobility disability; consequently, they may lack sufficient skeletal muscle function to allow for successful pelvic floor muscle training.[1, 7] We hypothesized that presence of impaired physical function may decrease the efficacy of pelvic floor muscle training in older women with urinary incontinence. The objective of this prospective study was to examine the effectiveness of behavioral and pelvic floor muscle training in older women with moderate-to-severe incontinence concomitant with physical function impairments.

## STUDY DESIGN, MATERIALS, AND METHODS

We conducted a prospective cohort study of 70 community-dwelling older women with at least moderate urinary incontinence symptoms. Institutional Review Board approval was obtained prior to study recruitment. Potential participants were identified using the electronic health record of women older than 70 years old with ICD-10 diagnosis codes for urinary incontinence within 6 months of the query [R32 (unspecified UI), N39.81 (functional UI), N39.41 (urge UI), N39.46 (mixed UI), and N39.3 (stress UI)]. Using batch mailings, potential participants were contacted with an introductory letter informing them of their eligibility to participate in this urinary incontinence treatment study. The letter also included the 6-item Questionnaire for Urinary Incontinence Diagnosis (QUID), validated to establish urinary incontinence as a diagnosis, distinguish incontinence type, and measure change in severity over time.[8] In addition, the study advertised to the community of patients enrolled in the newsletter registry that targets older adults interested in volunteering for research studies in aging. Women interested in participating either in response to the introductory letter or the newsletter were instructed to call our study coordinator for further screening.

A telephone screen was conducted by trained study coordinators to determine eligibility using scripted questions and the QUID questionnaire. The telephone script included standardized questions regarding the potential participants’ current physical and cognitive abilities, and physical activity. Eligible women were: age ≥ 70 years with a diagnosis of at least moderate incontinence symptoms based on the QUID subscale score for stress ≥4; urge ≥ 6; or total ≥ 10 [8]; willing and able to be compliant with pelvic floor muscle training and log adherence; and willing and able to undergo an extensive physical function evaluation. Women were excluded if they had any of the following: history of a surgical intervention for incontinence or hysterectomy within the past 12 months; diagnosis of pelvic organ prolapse beyond the hymenal ring; urogenital fistula; neurogenic overactive bladder (associated with a diagnosis of multiple sclerosis or stroke within past 12 months); incomplete bladder emptying or retention of urine greater than 150 ml (measured by bladder scan); required an assistive device (4-point cane, walker) for ambulation all or most of the time or wheelchair bound; undergoing treatment for significant cognitive impairment or dementia; unsafe to exercise (severe cardiopulmonary disease); unable/unwilling to sign informed consent; or if determined otherwise ineligible by the PI. All participants provided written informed consent to participate in the study according to the guidelines set forth by the Wake Forest School of Medicine. Eligible participants were invited for an in-person baseline visit. All study visits were conducted at the Geriatric Clinical Research Unit in our Claude D. Pepper Older Americans Independence Center.

Important clinical and demographic data were collected at baseline. Overall health status and 10-year mortality risk was assessed using the Charlson Co-morbidity Index. [9] The Montreal Cognitive Assessment (MoCA) was used to assess cognitive function.[10] The MoCA scoring ranges from 0-30. Scores between 19-25 define mild cognitive impairment. Alzheimer’s dementia is defined by score <21. Emotional health was assessed using the Center for Epidemiologic Studies Depression (CESD-10), a validated depression-screening tool that assesses depressive symptoms in the past week. A cut-off score of ≥10 represents significant depressive symptoms. [11]

Urinary incontinence and pelvic floor symptom assessment was performed through interview and validated questionnaires. A 3-day bladder diary established daily voiding and incontinence episodes collected at baseline, 6, and 12 weeks.[12] Based on the 3-day bladder diary, daytime voiding episodes were those occurring from the first void of the day to the last void prior to sleep and nighttime voiding episodes were those occurring after onset of sleep and before awakening. The mean total incontinence episodes were classified by the total number of stress and/or urgency episodes that occurred and were averaged over the 3-day diary. Urinary incontinence symptom impact on daily life was assessed using the Urinary Distress Inventory (UDI-6); scores greater than 33.3 indicate high distress from urinary incontinence symptoms.[13, 14] Bowel habits were assessed using the BRISTOL stool chart.

Pelvic examination and pelvic floor muscle assessment was performed by a Urogynecologist (PI) or a certified Pelvic floor Physical Therapist. Baseline pelvic floor muscle function was assessed objectively using the intravaginal Peritron® perineometer and the PERFECT scheme. PERFECT is an acronym used to ensure assessment of the main components of pelvic floor contractility[15]: P= power (a measure of maximal strength using vaginal digital exam or manometric perineometer), E=endurance (how long can women hold the contraction up to a max of 10 seconds), R=repetitions (how many maximal hold strength repetitions can women sustain up to 10 rep), F=fast contractions (how many fast maximal contractions can be repeated), ECT = every contraction timed (how long do women hold the fast contractions). Pelvic floor power was determined objectively using the Peritron® perineometer by obtaining a mean pressure measurement over 10 contractions of maximal effort.

The pelvic floor muscle training prescription for each participant was based on their individual PERFECT assessment and included 3-sets of daily pelvic floor exercises with each set followed by rapid contractions for 12 weeks. Based on physical therapy and exercise science literature, 8-12 repetitions of 3-4 sets over 6 weeks typically increases strength and efficiency of muscle recruitment.[16] Urgency and stress suppression strategies were included as appropriate to isolate the pelvic floor through rapid contractions when urinary urgency occurred, or a stress-provoked urinary incontinence episode was anticipated. Behavioral management strategies included recommendations for fluid and bowel management as appropriate.

The Short Physical Performance Battery (SPPB) was used to determine lower extremity physical function. The SPPB is a standard and robust predictor for disability that includes progressively more challenging standing balance tasks held for 10 seconds each (side-by-side, tandem, and semi-tandem), the faster of two 4-m courses at usual pace, and time to complete five repeated chair stands. [17, 18] Each of the three performance measures was assigned a score ranging from 0 (inability to perform the task) to 4 (the highest level of performance) and summed to create an SPPB score ranging from 0 to 12 (best). A low SPPB score (≤ 9) is a strong risk factor of decreased mobility and activities of daily living (ADL) disability in nondisabled older adults and was used in this study to define impaired physical function.[19]

Participants were asked to perform leg-extensions at 60 degrees per second on the isokinetic dynamometer (Biodex®) as an objective measure of maximal isokinetic lower extremity strength.[20] A whole-body DEXA scan was performed to determine appendicular lean muscle mass; the lean muscle mass index was used as an indicator for sarcopenia based on validated cut-offs.[21, 22]

### Statistical considerations

While the extent of the intervention extended through 12 weeks, our primary outcome was the change in urinary incontinence episodes from baseline to 6 weeks based on the presence of impaired physical function at baseline. In clinical practice, it is common to perform an assessment of urinary incontinence outcomes after 6 weeks of PFM training.[23] Therefore, we used this time point to strengthen the pragmatic impact of our observations. Secondarily, we examined the change in strength of pelvic floor muscles, the estimated global impression of improvement, and satisfaction rates at 6 and 12 weeks.

Baseline means and standard deviations for continuous variables and frequencies and percentages for categorical variables were used to describe the sample by physical function status. Tests of difference utilized Student’s T-test, Wilcoxon rank sum non-parametric test, or Chi-square for continuous and categorical measures, respectively. We estimated that a sample size of 60 women (30 per physical function group) would provide 82% power to detect a decrease of one urinary incontinence episode/day. Longitudinal mixed model linear regression analyses were used to assess for change in urinary incontinence episodes, overall satisfaction, global impression of improvement, and estimate of improvement based on the presence of physical function impairment adjusted for age, race, and BMI.

## RESULTS

Two-hundred fifty-three women were phone screened between January 2018 – January 2020. One-hundred and seventy-three women were ineligible on phone screening leaving 80 women who completed in-person baseline screening. Ten women screen-failed at the baseline visit leaving 70 women included in our analysis (Figure 1). At baseline, 33 women had impaired physical function (SBBP≤ 9) and 37 had normal physical function (Table 1). The mean age was 76.9 ± 5.4 years and our sample was 15.7% African American. There were no differences in mean age or race between physical function groups at baseline. Women with physical function impairment had a higher mean BMI than women without (P=0.032). Ten-year survival based on the mean±SD Charlson comorbidity index of 4.5±0.8 was approximately 50%. Depressive symptoms were uncommon. Mean MoCA scores for the cohort was 24.6 ± 3.0, indicating mild cognitive impairment was present in the 50% of women, with no differences based on physical function status. (Table 1)

**Figure 1.**
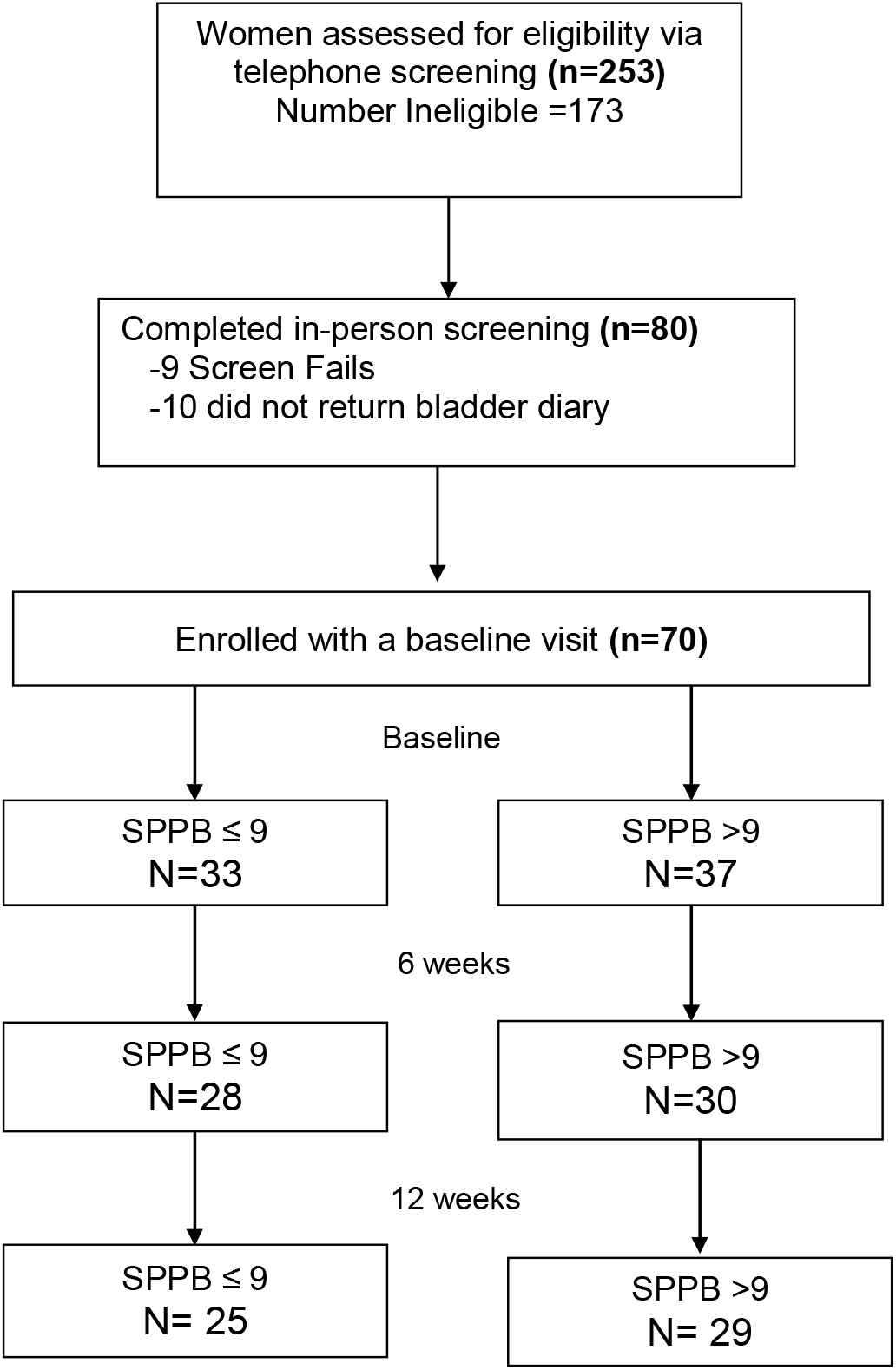
Flow diagram of patient identification via the electronic medical record, patient recruitment via telephone screening, and presentation for baseline evaluation.

**Table 1.**
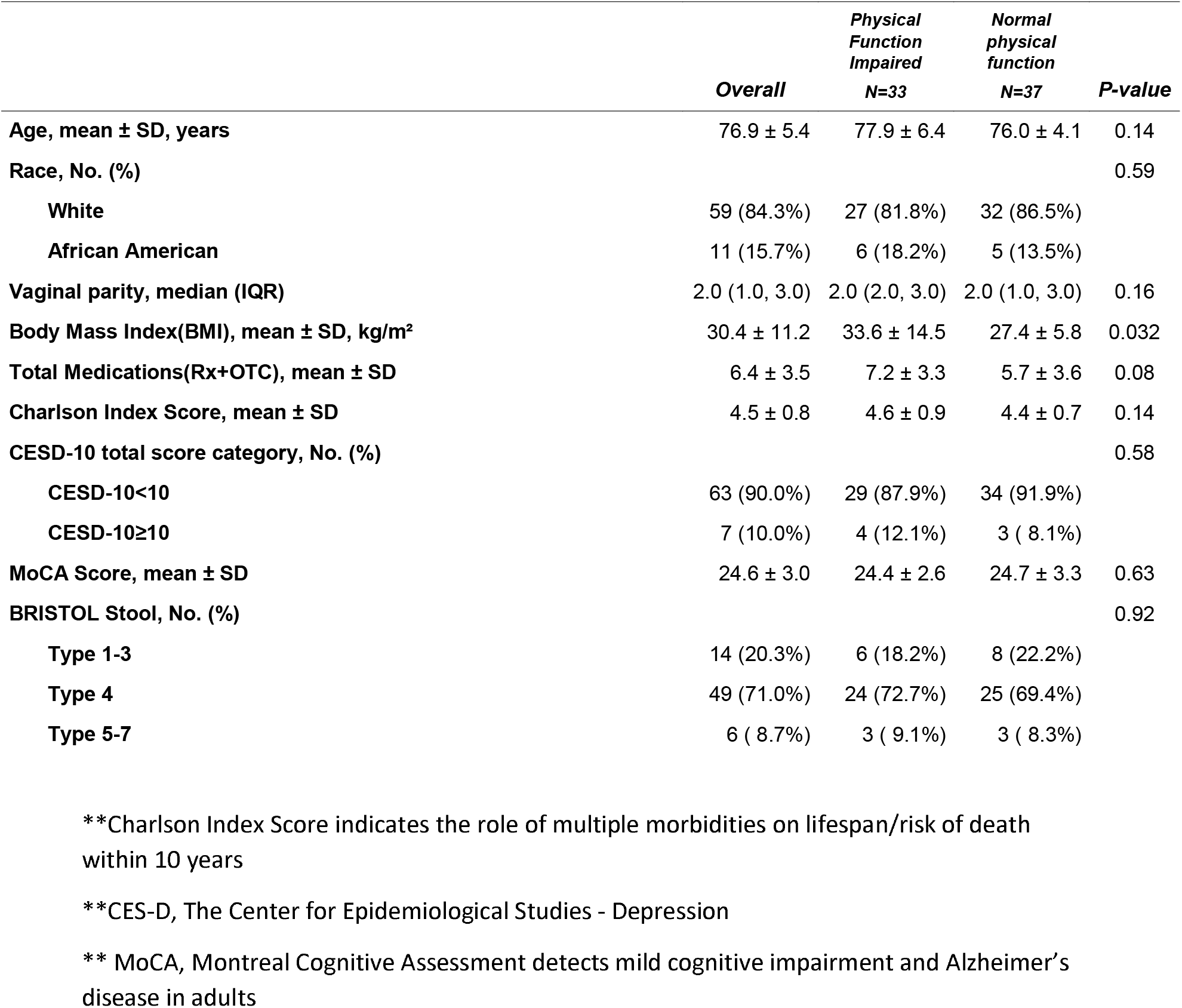
Demographic characteristics of the cohort by SPPB functional group.

Baseline daytime and nighttime mean voiding episodes were similar between groups (Table 2). Nocturia was common with 67.2% having nocturia (≥2 nights voids) with no difference based on functional groups. At baseline, women with physical function impairments had significantly more urinary incontinence episodes compared to women with normal physical function [4.5±2.9 vs. 2.7±2.1 UI episodes/day, p=0.0053]. Urgency urinary incontinence was predominant in women with physical function impairments compared to women without (p=0.037). Bother from urinary incontinence symptoms at baseline, as determined by the UDI-6 total scores, were low (mean±SD of the cohort, 16.7±4.9, p=0.42) and did not differ by physical function status. (Table 2) None of the validated questionnaires captured the significant differences that we observed in daily urinary incontinence episodes between groups according to bladder diary results.

**Table 2.** Pelvic Floor outcomes of the cohort by SPPB functional group.

|  | *Overall* | *Physical*<br>*Function*<br>*Impaired* | *Normal*<br>*physical*<br>*function* | *P-value* |
| --- | --- | --- | --- | --- |
| *Pelvic floor outcomes* |  |  |  |  |
| **3 day voiding diary, mean±SD** |  |  |  |  |
| Day time voiding episodes, mean ± SD, per day | 7.1 ± 2.9 | 7.3 ± 3.2 | 6.9 ± 2.7 | 0.64 |
| Night time voiding episodes, mean ± SD, per day | 2.5 ± 1.5 | 2.5 ± 1.8 | 2.5 ± 1.2 | 0.94 |
| Total UI episodes, mean ± SD, per day | 3.6 ± 2.7 | 4.5 ± 2.9 | 2.7 ± 2.1 | 0.0053 |
| **QUID** |  |  |  |  |
| Total Score, mean ± SD | 15.4 ± 4.8 | 15.6 ± 5.6 | 15.2 ± 4.0 | 0.76 |
| Stress Score, mean ± SD | 6.5 ± 3.5 | 6.8 ± 3.5 | 6.3 ± 3.5 | 0.53 |
| Urgency Score, mean ± SD | 9.1 ± 2.5 | 9.3 ± 3.2 | 8.9 ± 1.9 | 0.59 |
| Urinary Distress Index (UDI-6), mean ± SD | 16.7 ± 4.9 | 16.2 ± 4.5 | 17.1 ± 5.3 | 0.42 |
| Pelvic Floor Impact Questionnaire UIQ-7 Score, mean ± SD | 38.2 ± 25.6 | 40.9 ± 20.2 | 35.7 ± 29.6 | 0.39 |
| *Skeletal muscle health outcomes* |  |  |  |  |
| **PERFECT** |  |  |  |  |
| Peak cmH2O Grade Mean of 10 Trials, mean ± SD | 24.4 ± 15.5 | 19.2 ± 13.3 | 29.3 ± 16.0 | 0.0070 |
| Power based on BRINK grade, mean ± SD | 3.0 ± 1.0 | 2.8 ± 0.9 | 3.1 ± 1.0 | 0.24 |
| Endurance, mean ± SD | 4.6 ± 2.0 | 4.4 ± 2.0 | 4.7 ± 1.9 | 0.49 |
| Repetitions of Max Power, mean ± SD | 9.0 ± 1.9 | 9.1 ± 1.9 | 8.8 ± 1.9 | 0.53 |
| Number of Fast Contractions in 10 seconds, mean ± SD | 4.5 ± 1.6 | 4.3 ± 1.7 | 4.7 ± 1.5 | 0.32 |
| Appendicular Lean Mass Adjusted for BMI, mean ± SD, m<sup>2</sup> | 0.6 ± 0.1 | 0.5 ± 0.1 | 0.6 ± 0.1 | 0.0073 |
| Strength BIODEX, mean ± SD, kg | 72.3 ± 20.6 | 66.2 ± 23.4 | 77.0 ± 17.2 | 0.05 |

Skeletal muscle strength was globally weaker among women with physical function impairments as was baseline pelvic floor muscle maximal strength measured with the perineometer (19.2±13.3 cm H_2_0 vs 29.3±16.0 cm H_2_0, p=0.0070). Women with physical function impairments also had significantly lower appendicular lean muscle mass (p=0.0073) and significantly weaker lower extremities as indicated by BIODEX scores (p=0.05) than women with normal physical function (Table 2).

At 6 weeks, we retained 28/33 (85%) of women with physical function impairments and 30/37 (81%) with normal physical function (Figure 1). After 6 weeks of pelvic floor muscle training and behavioral therapy, women with physical function impairments experienced a non-significant decrease in urinary incontinence episodes (mean [95%CI]: −1.2[−2.0,−0.5] UI episodes/day] compared to women without physical function impairments (−0.4[−1.1, 0.3]) with no difference in mean change between groups (p=0.31). Women with physical function impairments also had a non-significant increase in PFM strength from baseline with mean peak strength over 10 trials being +2.2(−1.1, 5.6) cm H_2_0 compared to peak strength of −0.8(−4.1, 2.6) among women with normal physical function. These observed changes in pelvic floor muscle strength were not different between groups (p=0.75) (Table 3). Women with normal physical function reported a non-significant, but greater decrease in bother from their incontinence symptoms from baseline compared to women with impaired physical function (UDI-6 total scores, SPPB>9 −2.9[−4.5, −1.2] vs. SPPB≤9 −0.6 [−2.4, 1.1], p=0.75).

**Table 3.** Examining the impact of poor physical performance (defined as SPPB>9) on reduction of UI episodes after 6 and 12 weeks of pelvic floor physical therapy from longitudinal mixed models adjusting for age, BMI, and race in addition to correlation between repeated measures.

|  | **Baseline**<br>**Mean±SE** |  |  | **Change at 6 weeks**<br>**mean(95%CI)** |  |  |  | **Change at 12 weeks**<br>**mean(95%CI)** |  |  |  | **SPPB\*Time**<br>**P-value** |
| --- | --- | --- | --- | --- | --- | --- | --- | --- | --- | --- | --- | --- |
|  | *Physical*<br>*Function*<br>*Impaired* | *Normal*<br>*physical*<br>*function* | *P-*<br>*value* | *Physical*<br>*Function*<br>*Impaired* | *Normal*<br>*physical*<br>*function* | *Diff* | *P-*<br>*value* | *Physical*<br>*Function*<br>*Impaired* | *Normal*<br>*physical*<br>*function* | *Diff* | *P-*<br>*value* |  |
| **3 day voiding**<br>**diary, mean±SD** |  |  |  |  |  |  |  |  |  |  |  |  |
| **Total leaks** | 4.3±0.6 | 2.7±0.6 | **0.0278** | -1.2<br>(-2.0,-0.5) | -0.4<br>(-1.1,0.3) | 0.8<br>(-0.7,2.2) | 0.31 | -0.8<br>(-1.6,+0.0) | -0.2<br>(-1.0,0.5) | 1.1<br>(-0.4,2.6) | 0.15 | 0.28 |
| **Urgency with**<br>**voids** | 1.1±0.3 | 1.9±0.3 | **0.0370** | 0.3<br>(-0.3,0.8) | -0.6<br>(-1.1,-0.1) | 0.0<br>(-0.7,0.8) | 0.90 | -0.3<br>(-0.8,0.3) | -0.6<br>(-1.1,-0.1) | -0.4<br>(-1.2,0.3) | 0.28 | 0.07 |
| **UDI-6 score,**<br>**mean±SD** | 15.7±1.1 | 17.4±1.1 | 0.20 | -0.6<br>(-2.4,1.1) | -2.9<br>(-4.5,-1.2) | 0.5<br>(-2.4,3.4) | 0.75 | -1.1<br>(-2.9,0.8) | -3.7<br>(-5.4,-2.1) | 0.9<br>(-2.1,3.9) | 0.55 | 0.07 |
| **Peak Pelvic**<br>**Floor muscle**<br>**strength, mean**<br>**± SD** | 21.1±3.3 | 27.5±3.3 | 0.12 | 2.2<br>(-1.1,5.6) | -0.8<br>(-4.1,2.6) | -3.5<br>(-12.0,5.0) | 0.42 | 1.9<br>(-1.7,5.4) | -0.9<br>(-4.4,2.7) | -3.7<br>(-12.3,4.8) | 0.39 | 0.39 |

After 12 weeks of pelvic floor muscle training, we retained 25/33 (76%) of women with baseline physical function impairment and 29/37 (78%) with normal physical function. Observations in change in incontinence episodes remained consistent. Women with physical function impairments had a small-sustained decrease in their incontinence episodes from baseline to 12 weeks; though this decrease was not significantly different compared to women with normal physical function (Table 3). After 12 weeks of training, women with impaired physical function had a greater increase in pelvic floor muscle strength from baseline with mean peak strength over 10 trials being +1.9(−1.7, 5.4) cm H_2_0 compared to −0.9(−4.4, 2.7) than women with normal physical function (p=0.21).

The majority (93%) of women with normal physical function self-reported being better or much better after 6 weeks of pelvic floor muscle training and behavioral therapy compared to only 69% of women with impaired physical function (Table 4, p=0.03). Overall, complete satisfaction with incontinence symptom improvement was low between both groups with of women; 41.8% of women with physical function impairments and 44.8% of women without physical function impairments were completely satisfied after 12 weeks of pelvic floor muscle training., p=0.02. Women with normal physical function had 2.8 higher odds of rating their impression of improvement as better/much better compared to women with physical function impairments. Differences in estimates of improvement were significantly lower among women with physical function impairments compared to women with normal physical function (Table 4).

**Table 4.** Examining satisfaction and improvement rates regarding reduction of UI episodes after 6 and 12 weeks of pelvic floor physical therapy from longitudinal mixed models adjusting for age, BMI, and race in addition to correlation between repeated measures.

|  | Subjective improvement/satisfaction @ 6 weeks |  |  |  | Subjective improvement/satisfaction @ 12 weeks |  |  |  | SPPB*Time<br>P-value |
| --- | --- | --- | --- | --- | --- | --- | --- | --- | --- |
|  | *Physical*<br>*Function*<br>*Impaired* | *Normal*<br>*physical*<br>*function* | *Normal vs physical*<br>*function impaired*<br>*OR(95%CI)* | *P-*<br>*value^* | *Physical*<br>*Function*<br>*Impaired* | *Normal*<br>*physical*<br>*function* | *Normal vs*<br>*physical function*<br>*impaired*<br>*OR(95%CI)* | *P-*<br>*value^* |  |
| Overall satisfaction |  |  |  | 0.7852 |  |  |  | 0.92 | 0.92 |
| Completely<br>N(%) | 10(38.5%) | 13(44.8%) | 1.0<br>(0.3,3.3)<br>p=0.90 |  | 10(41.8%) | 13(44.8%) | 1.1<br>(0.3,3.6)<br>p=0.97 |  |  |
| Not at<br>all/Somewhat N(%) | 16(61.5%) | 16(55.2%) | REF |  | 14(58.3%) | 16(55.2%) | REF |  |  |
| Global impression<br>of improvement |  |  |  | 0.0346 |  |  |  | 0.12 | 0.78 |
| Better/Much<br>better N(%) | 18(69.2%) | 27(93.1%) | 2.8<br>(0.5,16.6)<br>p=0.25 |  | 18(75.0%) | 27(93.1%) | 4.1<br>(0.8,20.3)<br>p=0.08 |  |  |
| About the same or<br>Worse/Much<br>worse N(%) | 8(30.8%) | 2(6.9%) | REF |  | 6(25.0%) | 2(6.9%) | REF |  |  |
| Estimate of<br>improvement (%) | *Mean±SE* | *Mean±SE* | *Mean diff (95%CI)* |  | *Mean±SE* | *Mean±SE* | *Mean diff (95%CI)* |  |  |
|  | 50.7±5.9 | 64.2±5.9 | 13.5<br>(-1.8, 28.9) | 0.08 | 49.7±5.9 | 64.2±5.9 | 18.0<br>(2.5, 33.5) | 0.0238 | 0.51 |

## DISCUSSION

In this study, the efficacy of pelvic floor muscle training, a primary recommended intervention strategy for urinary incontinence, did not provide a significant reduction in urinary incontinence symptoms or satisfaction for older women with moderate to severe incontinence symptoms, regardless of baseline global physical function impairment. Our data strengthen observed associations between physical function impairments and urinary incontinence severity. Among older women with physical function impairment at baseline, small (−1.3 episodes) improvements in incontinence symptoms were achieved by 6 weeks and sustained to 12 weeks with pelvic floor muscle training and behavioral therapy. This decrease in incontinence episodes/day did not result in minimum clinically important improvements in UDI-6 scores.Thus, despite reports of “being better”, bother and distress from their incontinence episodes remained. The high percentage of women who reported a favorable global impression of improvement may reflect a desire to appease the study staff regarding the intervention. From our cohort, we did not confirm our hypothesis that poor baseline physical function corresponded with major differences in pelvic floor exercise responsiveness.

While meta-analyses have confirmed the utility of pelvic floor muscle exercises for all types of urinary incontinence compared to placebo, Cammu et al identified several key predictors of treatment failure most notably of which was baseline severity of incontinence (>2 episodes per day).[24, 25] It is plausible that we selected a population of women that had crossed the threshold of severity for which pelvic floor muscle therapy might be efficacious, regardless of age or global physical function. Baseline pelvic floor muscle strength and global physical function have not previously been considered when recommending pelvic floor muscle training to older incontinent women. In our cohort, we noted a strong corollary between pelvic floor strength and global physical function performance metrics. This may reflect a global weakness in skeletal muscle as evidenced by lower appendicular lean mass, weaker lower extremities, and higher incidence of sarcopenia among women with concomitant physical function impairment.[1] Contradictory to our stated hypothesis, however, women with poor overall physical function actually demonstrated a trend towards greater improvement in their pelvic floor strength after directed pelvic floor muscle training than women with normal global physical function. Therefore, this low-risk intervention may remain a reasonable treatment option in elderly, frail women especially if it is introduced before symptoms advance in severity.

Several studies have affirmed the association between poor physical function and urinary incontinence. Tinetti et al studied a cohort of older community-dwelling adults to assess predisposing factors associated with urinary incontinence, falling, and functional dependence. Lower extremity weakness (poor performance on timed chair stands) showed the strongest relationship with these factors.[2] In a secondary cross-sectional analysis among women with daily urinary incontinence, 24% reported specific difficulty or dependence with using the toilet, and were 3.3 times more likely to have functional difficulty or dependence compared to continent older women.[26] [27] Nearly a quarter of women older than 65 years who participated in the California Health Interview Survey reported symptoms; this was significantly associated with poorer overall health (adjusted OR 3.43), decreased mobility (OR, 1.81), and history of falls (OR, 1.53).[28] Current practice guidelines should consider these important age-related changes when considering incontinence treatment.[29]

The greatest strength of this work is in the population studied. Women with urinary incontinence who are older than 70 years are rarely included in clinical trials. In addition, the comprehensive evaluation for physical function impairments is a novel occurrence in clinical trials designed to observe effectiveness of non-surgical treatment for urinary incontinence. Our findings are strengthened by our use of validated measures to determine changes in urinary incontinence episodes, pelvic floor muscle strength, and for assessment of satisfaction with treatment. The use of subjective and objective outcome measures for urinary incontinence further strengthens this work as it provides a more wholistic assessment of the impact of physical function impairments on the success of pelvic floor muscle exercise to treat urinary incontinence in older women. Lastly, we were able to successfully recruit 70 women older than 70 years into this prospective study with a retention rate of 77% at 12 weeks.

While robust in design and implementation, there are several important limitations to this study that may influence the conclusions. First, our criterion for incontinence severity upon enrollment targeted women with at least moderate symptoms. It is plausible that the impact of pelvic floor muscle exercises may only be in women with mild symptoms; thus explaining the lack of effectiveness observed in this cohort. Had we chosen women with mild incontinence, it is plausible that pelvic floor muscle exercises could have resulted in significant improvement and satisfaction, particularly in the absence of physical function impairment. However, we previosly documented that greater severity of urinary incontinence is associated with physical function impairments.[6] Therefore, targeting women with more severe symptoms to understand the role of pelvic floor muscle training in this cohort of understudied women was the purpose of this work. A criticism to our findings is in the pragmatic design of the intervention implementation. The majority of older women with urinary incontinence symptoms seeking treatment are not routinely referred to pelvic floor physical therapy for the conduct of pelvic floor muscle training as the first-line treatment for their urinary incontinence symptoms. Access to trained pelvic floor physical therapy is limited by a cost, location of certified providers, and the sacrifice of appointment times. However, supervised pelvic floor muscle training by a certified pelvic floor physical therapist may have superior results than home prescriptions as used in this study. Our pragmatic design of progressive, home-based pelvic floor training and behavioral therapy is significantly more generalizable compared to a more supervised approach.

In conclusion, ageing and physical function impairment strongly affect urinary incontinence rates and conservative behavioral and pelvic floor muscle interventions appear to have limited impact in this population. The demonstrated positive impact on pelvic floor muscle strength, however, even in women with poor global physical function, raises the question of utilization of these exercises as a more effective prevention than treatment strategy. It is also possible that pelvic floor muscle exercises could be an important intervention for limiting progression of incontinence severity in women with declining physical function. However, advancements in non-surgical treatments are needed to more significantly influence urinary incontinence severity and to improve treatment satisfaction for older incontinent women.

## Data Availability

All data produced in the present study are available upon reasonable request to the authors

